# Fake Publications in Biomedical Science: Red-flagging Method Indicates Mass Production

**DOI:** 10.1101/2023.05.06.23289563

**Authors:** Bernhard A. Sabel, Emely Knaack, Gerd Gigerenzer, Mirela Bilc

## Abstract

**Background:** Integrity of academic publishing is increasingly undermined by fake science publications massively produced by commercial “editing services” (so-called “paper mills”). They use AI-supported, automated production techniques at scale and sell fake publications to students, scientists, and physicians under pressure to advance their careers. Because the scale of fake publications in biomedicine is unknown, we developed a simple method to red-flag them and estimate their number.

**Methods:** To identify indicators able to red-flag fake publications (RFPs), we sent questionnaires to authors. Based on author responses, a classification rule was applied initially using the two-indicators “non-institutional email AND no international authors” (“email+NIA”) to sub-samples of 15,120 PubMed®-listed publications regarding publication date, journal, impact factor, country and RFP citations. Using the indicator “hospital affiliation” (“email+hospital”), this classification (tallying) rule was validated by comparing 400 known fakes with 400 matched presumed non-fakes.

**Results:** Two initial indicators (“email+NIA”) revealed a rapid rise of RFP from 2010 to 2020. Countries with the highest RFP proportion were Russia, Turkey, China, Egypt, India and China (39%-55%). When using the “email+hospital” tallying-rule, sensitivity of RFP identification was 86%, the false alarm rate 44%, and the estimated RFP rate in 2020 was 11.0%. Adding a RFP-citation indicator (“email+hospital+RFP-citations”) increased the sensitivity to 90% and reduced the false alarm rate to 37%. Given 1.3 million biomedical Scimago-listed publications, the estimated annual RFP number in 2020 is about 150,000.

**Conclusions:** Potential fake publications can be red-flagged using simple-to-use, validated classification rules to earmark them for subsequent scrutiny. RFP rates are increasing, suggesting higher actual fake rates than previously reported. The large scale and proliferation of fake publications in biomedicine can damage trust in science, endanger public health, and impact economic spending and security. Easy-to-apply fake detection methods, as proposed here, or more complex automated methods can enable the retraction of fake publications at scale and help prevent further damage to the permanent scientific record.

## INTRODUCTION

Trust in the integrity of academic publishing is a foundation of science, and lack of it damages its reputation (Behl, 2021; Byrne, 2019; Seifert, 2021; Else & Van Norden, 2021; Else, 2022; Byrne et al., 2022). Well-known cases of scientific misconduct by individual researchers include ghost and “honorary” authorships (Flanagin et al., 1998; Wislar et al., 2011; Frederickson and Herzog, 2021), cherry-picking, abstract spin, plagiarism of images (Bik et al., 2016), and outright data fabrication (Bik, 2020; Byrne and Christopher, 2020; Park et al., 2022). While individual fraud has been recognized for centuries, the recent emergence of commercial production of fake publications is a new and unprecedented development (Flanagin et al, 1998; Wislar et al., 2011; Mavrogenis et al., 2018; Byrne, 2019; Byrne and Christopher, 2020; Else and Van Norden, 2021; Sabel and Seifert, 2021; Sabel, 2022; Chawla Singh, 2022; Candal-Pedreira et al., 2022). The major source of fake publications are 1,000+ “academic support” agencies – so-called “paper mills“ – located mainly in China, India, Russia, UK, and USA (Abalkina, 2021; Else, 2021; Pérez-Neri et al., 2022). Paper mills advertise writing and editing services via the internet and charge hefty fees to produce and publish fake articles in journals listed in the *Science Citation Index* (SCI) (Christopher, 2021; Else, 2022). Their services include manuscript production based on fabricated data, figures, tables, and text semi-automatically generated using artificial intelligence (AI). Manuscripts are subsequently edited by an army of scientifically trained professionals and ghostwriters. Although their quality is still relatively low (Cabanac and Labbé, 2021), fake publications nevertheless often pass peer review in established journals with low to medium impact factors (IF 1-6) (Seifert, 2021). Some governments, funding bodies, and academic publishers are on the alert (Cyranoski, 2018; Mallapaty, 2020; Else, 2022; Candal-Pedreira et al., 2022), yet many scientists, journal editors, and learned societies appear to be surprisingly unaware that such publications exist at all.

Paper mill customers – students and scientists – are pressured to publish in SCI publications by their academic or government institutions or university-affiliated hospitals (Pérez-Neri et al., 2022). For example, the Beijing municipal health authorities require a fixed number of first-authored SCI articles for physicians to qualify for promotion (Else and Van Norden, 2021). Academic policies that count publications and value impact factors as surrogates for scientific excellence can force graduate students to fulfill SCI publication requirements and pressure scientists and physicians to meet publication quotas to attain salary increases, promotion, and/or scientific reputation. Paper mills are ready to help and offer their services to accomplish these goals.

We are aware of several instances where paper mills tried to promote their business by inviting journal editors to collaborate, as shown by this unsolicited email in 2022 from a paper mill to one of us who is editor of a biomedical journal (see Tab. 1 for interview excerpts):

> We are a well-known academic support institution from Guangzhou, China, which has been established for 8 years. … For reducing the publication time, we expect to cooperate with you in the future.

> Cooperation mode: we cite the content of your journal in our articles, thus increasing … your impact factor in 2022. You shall help us shorten the publication time. Payment: If an article is successfully published, we will pay for it at the price: IF*1,000 USD/article. For example, with

> IF=2.36, total payment=2.36*1,000 USD=2,360 USD. And this price is negotiable.

**Table 1:**
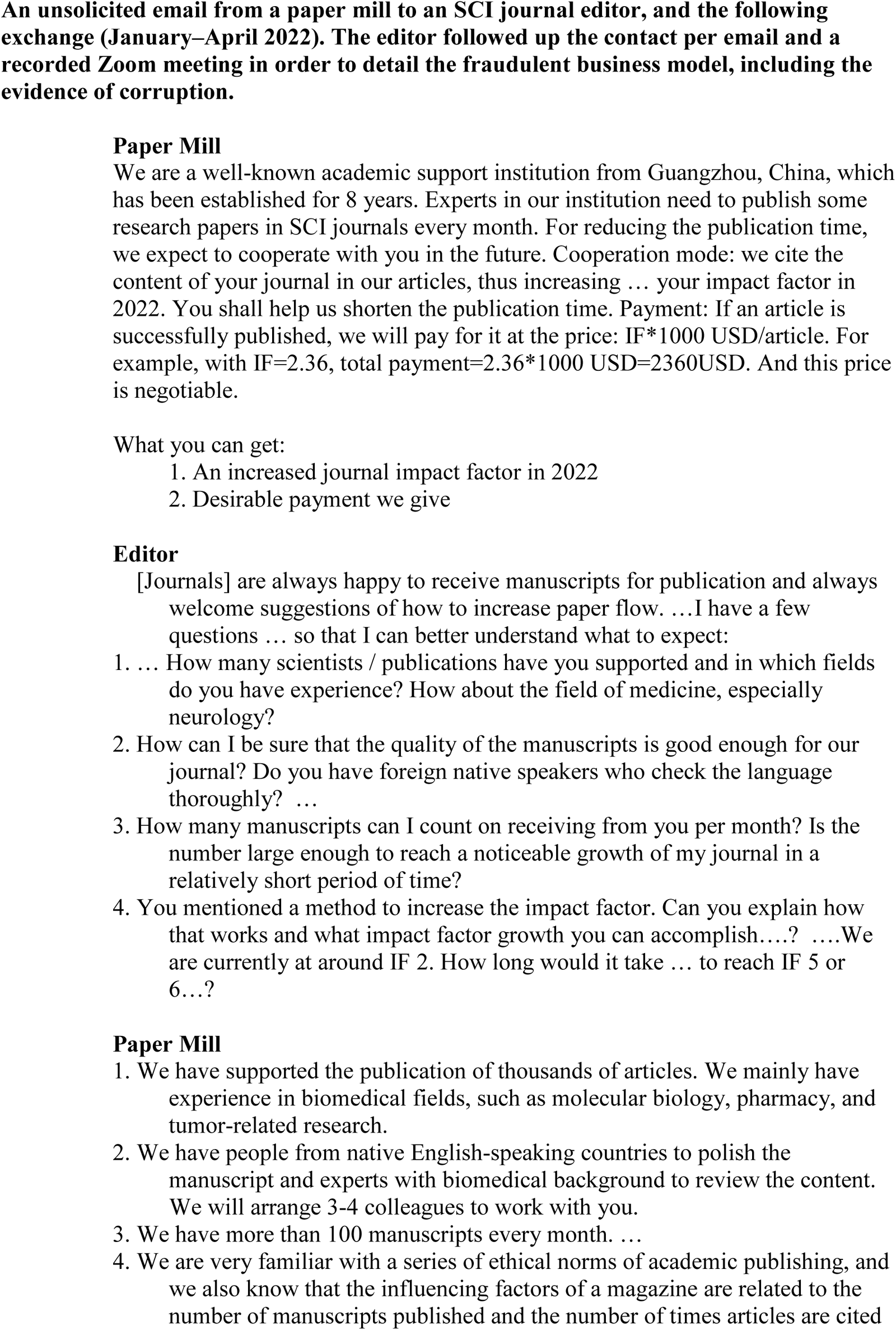
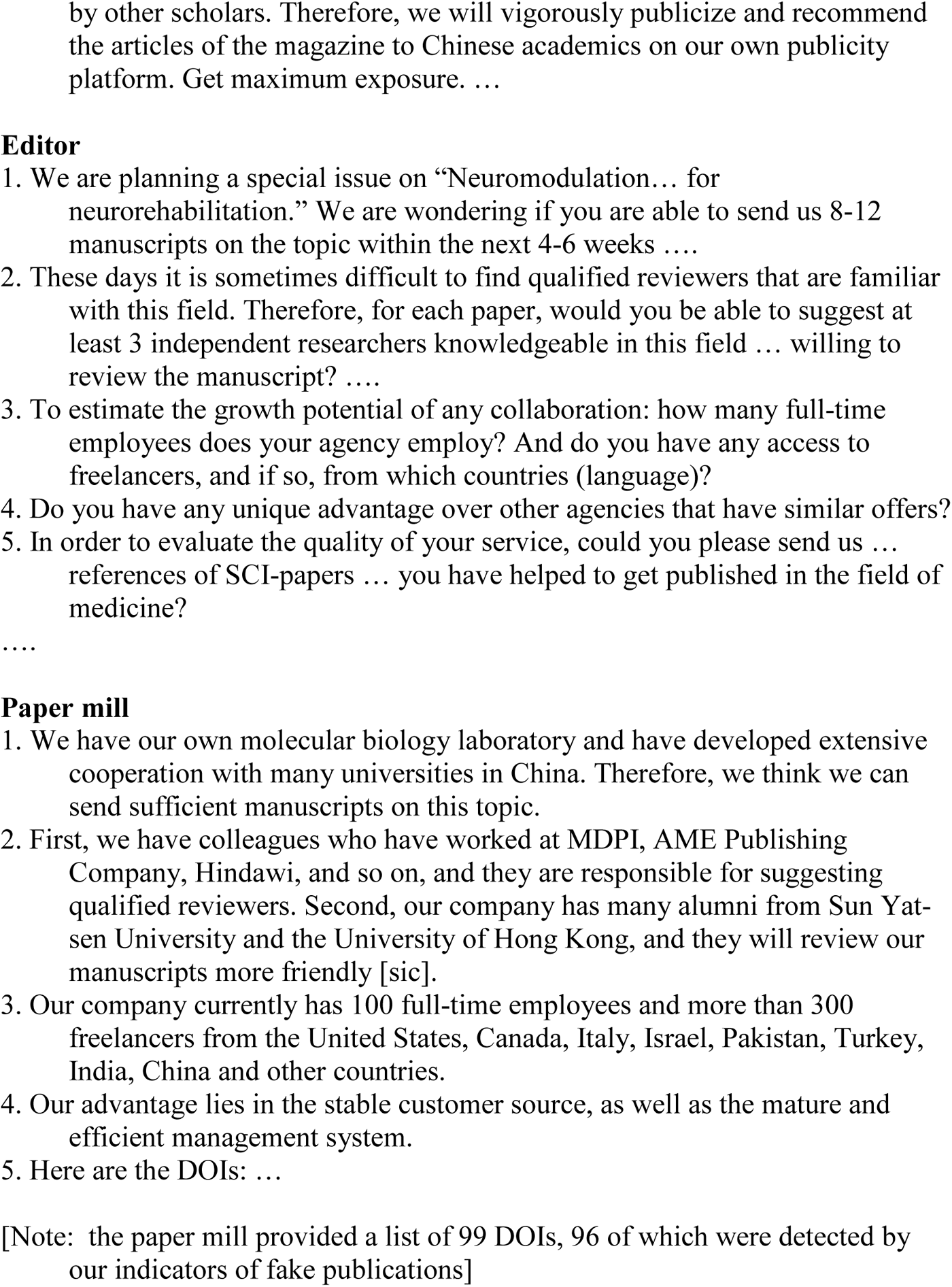
Rendezvous with a paper mill – A true story.

This attempted corruption motivated us to quantitatively analyze the global scope of fake publishing. Because the problem is still perceived to be small (an estimated 1 of 10,000 publications, Tab. 2), publishers and learned societies are just beginning to adjust editorial, peer-review, and publishing procedures. Yet the actual scale of fake publishing remains unknown, despite the fact that the number of reports on paper mills are increasing.

**Table 2:** Number of known fake publications in the biomedical literature.

| **Published reports of known fake publications** |  |  |  |
| --- | --- | --- | --- |
| **Reference** | **Yrs. analyzed** | **No. of fakes** | **Type of analysis** |
| Carlisle (2021) | 2007 - 2020 | 43 | baseline data analysis |
| Fisher and Cox (2021) | 2020 | 68 | suspected fraudulent papers |
| Heck et al (2021) | 2021 | 75 | image forensics, raw data requests |
| Seifert (2021) | 2019 - 2020 | 100 | raw data requests |
| Nath et al (2006) | 1982 - 2002 | 107 | retraction analysis |
| Mallapaty (2020) | 2017 | 107 | retraction analysis |
| Behl (2021) | 2017 - 2021 | 137 | image forensics |
| Elango (2021) | 1992 - 2020 | 161 | retraction analysis |
| Bozzo et al (2017) | 1946 - 2015 | 163 | retraction analysis |
| Lei et al (2018) | 1997 - 2016 | 260 | retraction analysis |
| Abalkina (2021) | 2019 - 2021 | 434 | offers from paper mill website |
| Bik (2020) | 2016 - 2020 | 633 | image forensics |
| Bik et al (2016) | 1995 - 2014 | 784 | image forensics |
| Brown et al (2022) | 2019 | 1,396 | retraction analysis |
| Fang et al (2012) | 2010 | 1,611 | retraction analysis |
| Candal-Pedreira | 2022 | 3,544 | retraction analysis |
| Ataie-Ashtiani (2018) | 2011 - 2017 | 4,931 | retraction analysis |
| **Total** |  | **14,554** |  |

*Note:* Examples of prior studies that quantified the number of fake publications in the years 1982-2022 as identified by text or image plagiarism, data fabrication, paper mill offers, retractions, etc. (Note: duplicate counts could not be determined). Several of these references cite additional publications, and 96.8% of paper mill retractions come from Chinese institutions (Candal-Pedreira et al., 2022). While our list is not exhaustive, given that the total number of biomedical publications is >15 million in the last decade, the percentage of currently identified actual fake publications is on the order of 0.1% for this time period compared with our estimate of 28% in 2020 alone.

To be able to estimate the scope of fake publishing, a method is needed to identify potential (red-flag) fake publications (RFPs). We therefore looked for potential indicators of fakes that are easy to use by reviewers, editors, and publishers and tested their feasibility for randomly selected neuroscience and medical journals. We then developed classification (tallying) rules for screening for potential fakes and determined their sensitivity and false alarm rates to estimate their global scope.

## METHODS

### Exploration

To search for potential fake indicators, in Study 1, one of us (editor of a biomedical journal) sent a questionnaire to the corresponding authors of a sample of suspicious published articles and, for control, to those of a sample of unsuspicious articles (see Tab.3). Based on the differential willingness to respond, we identified potential indicators for fake publications. These indicators should satisfy the following patterns (see below):

i. Authors of fake publications are reluctant to provide critical information as revealed by their response – or non-response – to the questionnaire by the editor,
ii. the number of fake publications increases steadily over time, and
iii. journals with a low to medium impact factor are most affected.

Based on our experience in Study 1, two easy-to-detect indicators were used in Studies 2-6 where publications were labelled as RFP if the corresponding author used a non-institutional (private) email with no international co-author (“email-NIA”). This indicator was applied to 15,120 publications listed in PubMed® and Web of Science™ in the fields of neuroscience (including neurology) and general medicine publications in a series of 9 bibliographic studies (Fig.1). Study 2 applied the email-NIA indicator to five neuroscience publications; Study 3 increased sample size to estimate RFP growth from 2010-2020, including five journals in the field of general medicine, Studies 4 and 5 we increased sample size further and Study 6 checked RFP rates in three open access journals (of the “Frontiers”-series).

**Figure 1.**
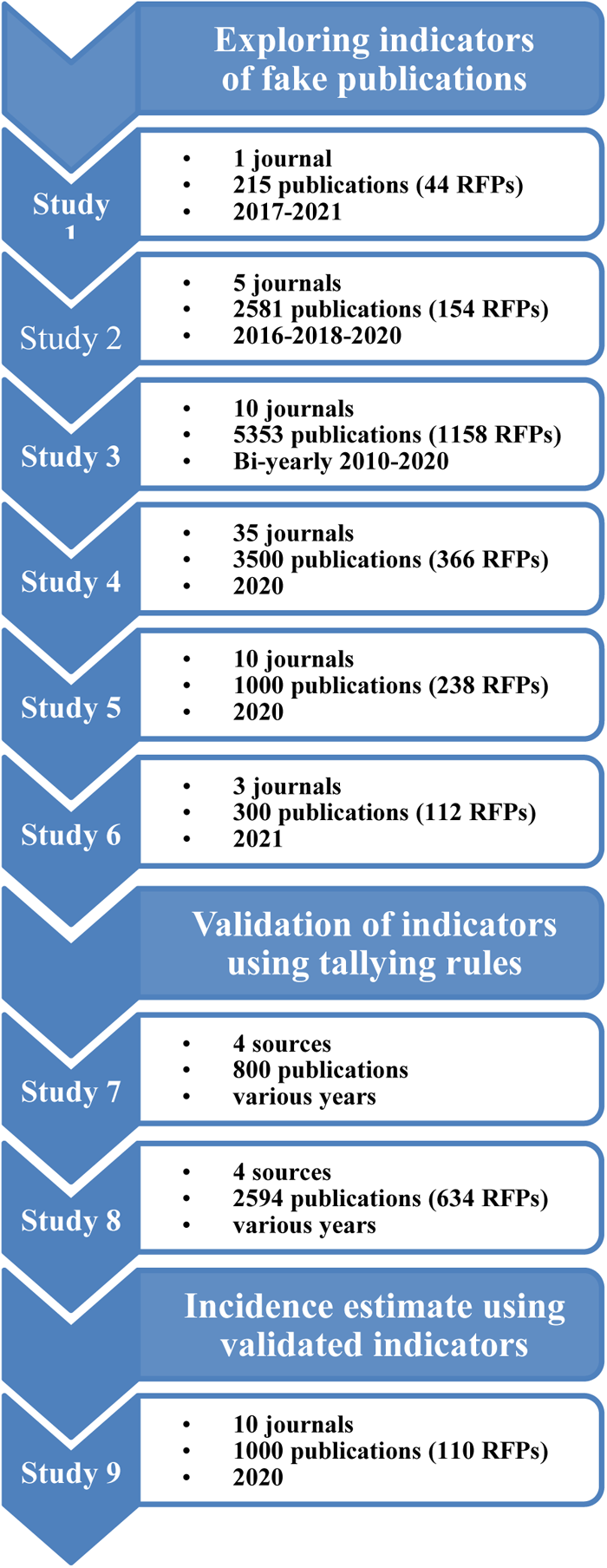
Screening plan for journals and publications. Studies 1-6 identified potential indicators of fake publications on the basis of hypotheses 1 to 3 (see text) using the indicators “private email” and “no international author”, Studies 7 used the indicator combination “email” and “hospital” as a feasible classification (tallying) rule with high sensitivity and acceptable false-alarm rate which was the applied to estimate the incidence of potential fake publications in Study 9. Study 8 was a test to check if the precision of the tallying rule of Study 7 can be increased by adding a third indicator (“RFP citation”). Note, some publications were used in several screening studies. The total number of unique publications analysed is n=15,120.

### Validation

Feasibility of additional indicators was checked in Study 7 by applying various tallying rules to a sample of n=400 “true fake” publications characterized by either fake gene sequences, text or image plagiarism, or if they were retracted:

i. n=100 retractions (using the criterion “paper mill”) (http://retractiondatabase.org/RetractionSearch.aspx?)
ii. n=100 Tadpole paper mill items (https://docs.google.com/spreadsheets/d/1KXqTAyl4j-jVorFPMD2XRpr76LcIKJ0CVyIvRj0exYQ/edit#gid=0)
iii. n=100 fake gene sequences; https://dbrech.irit.fr/pls/apex/f?p=9999:28
iv. n=100 retractions from *Journal of Cellular Biochemistry* (Behl, 2021).

The “true fake” sample was then matched with a n=400 presumed “non-fake” publications sampled from the same journals by selecting a fake publication’s nearest-neighbor article (unless it was also a known fake). A limitation of this “matched sample” method is that we cannot be certain whether the publications presumed “non-fake” may not actually be fake as well. The tallying-rule “email+hospital” was found to be the most feasible in view of its efficiency, sensitivity and acceptable false-alarm rate. To check if the precision of RFP detection can be increased, Study 8 added an additional indicator: percentage of RFPs citations (“email+hospital+RFP citations>10%”) in each of 80 randomly selected publications (40 proven fake and 40 presumed non-fake) (i.e., 2,594 additional publications).

### Estimating RFP incidences

To estimate the rate of RFPs in the biomedical literature, the Study 5 sample was re-analyzed in Study 9 using the validated “email+hospital” rule.

Finally, we studied >1,000 websites retrieved from Baidu and Google that advertise various editing services (search terms: “SCI-publication or –service,” “essay writing service,” “journal writing service,” “SCI ghost writing”) and interviewed a paper mill manager.

## RESULTS

### EXPLORATION OF FAKE INDICATORS

We searched for indicators that can be determined easily, quickly, and reliably by an editor on the basis of a submitted manuscript or publication alone. In an exploration phase (Study 1-6), our search was guided by three hypotheses:

#### Hypothesis 1: Authors of fake publications are unwilling to answer quality check surveys nor provide original data

In Study 1, n=215 neurology articles were manually inspected by an experienced editor; 20.5% (n=44) were deemed suspicious. A questionnaire was sent to all authors and, for control, to 48 authors of non-suspicious papers. It contained questions that authors of fake papers might be reluctant to answer (e.g., “Are you willing to provide original data?” [only 1 author of 44 suspicious articles did] and “Did you engage a professional agency to help write your paper? [none did]; see Tab. 3). Despite repeated reminders with a warning that failure to reply – or replying inadequately – could trigger retraction, the response rate among suspected authors was only 45.4% (20/44) compared with 95.8% (46/48) for the control group (24/215 non-responses). This survey provided the preliminary indicator (email+NIA) to red-flag publications (RFP).

**Table 3.**
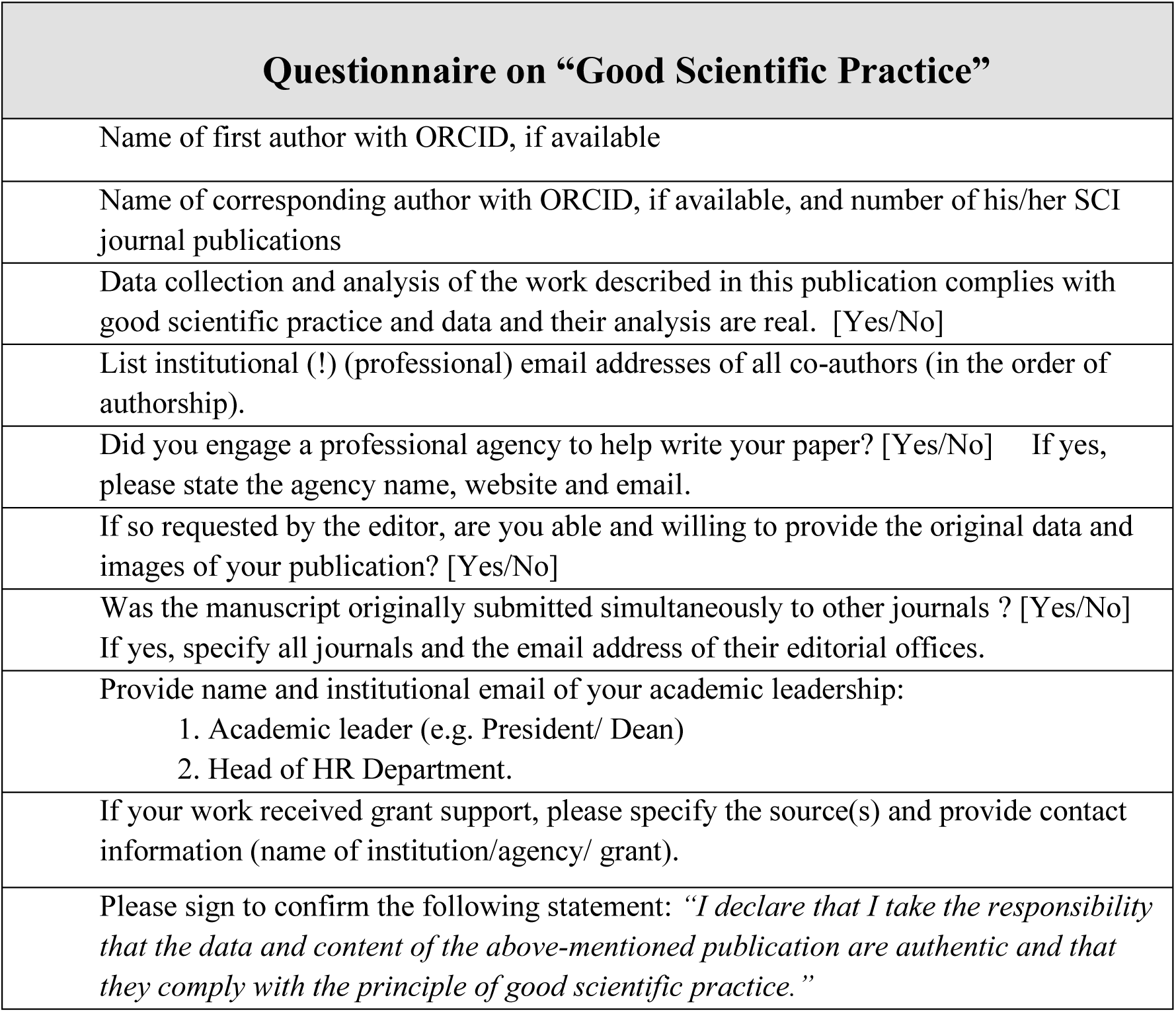
Questionnaire sent to corresponding authors.

#### Hypothesis 2: Because paper mills are on the rise (Else and Van Norden, 2021), indicators uncovered in Study 1, if valid, should also increase each year

Study 2 analyzed the frequency of these indicators in five randomly chosen neuroscience journals, expanded in Study 3 to a larger sample of articles from those five neuroscience journals and an additional five medical journals bi-annually (2010-2020). The results show a rapid growth of RFPs over time in neuroscience (13.4% to 33.7%) and a somewhat smaller and more recent increase in medicine (19.4% to 24%) (Fig. 2). A cause of the greater rise of neuroscience RFPs may be that fake experiments in basic science (biochemical-, *in vitro-* and *in vivo-*studies) are easier to generate because they do not require clinical trial ethics approval by regulatory authorities.

**Figure 2.**
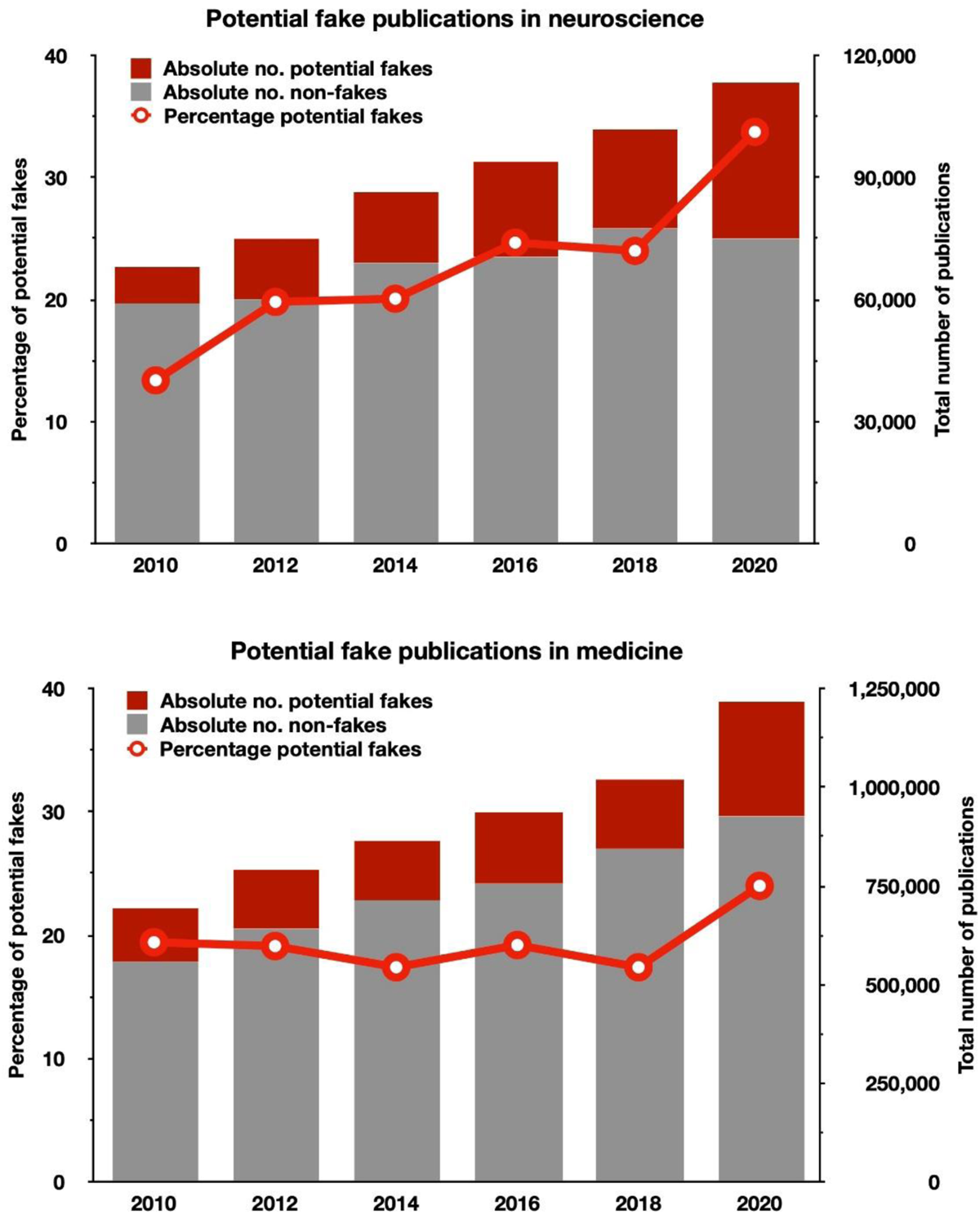
The rise of red-flagged fake publications (RFPs). Number of publications and percentage of red-flagged fake publications in medicine (top) and neuroscience (bottom) (Study 3). The red line (y1-axis) shows the percentage of red-flagged publications per year; the grey bars (y2-axis) show the estimates of non-fakes and the red bars those of potential fake publications, extrapolated to overall articles published in neuroscience (selected from journals of the Neuroscience Peer Review Consortium) and medicine (selected from scimagojr.com). Note the relatively slow increase in potential fakes in medicine and the rapid rise in neuroscience.

#### Hypothesis 3: Because it is easier for paper mills to market their papers to journals with lower impact factors (IF; range 1-6), our indicators, if valid, should also occur more frequently in such journals

Study 4 tested an even larger sample of randomly selected NPRC-Neuroscience journals which red-flagged 366/3,500 (10.5%) RFPs. In Study 5 counted RFPs in 10 randomly chosen general medical journals, where the 2020 rate was 23.8% in lower IF journals which was only topped by open access (OA) journals in Study 6 (112/300=40.3%).

### VALIDATION OF FAKE INDICATORS

To explore which fake indicators can best identify true fakes, we computed sensitivity/false alarm rates by comparing a sample of known fakes (n=400) with a sample of presumed non-fakes (n=400) combining the two best indicators (email+hospital) in a classification (tallying) rule: “If both indicators are present, classify as a potential fake, otherwise not” (the “AND” rule) (Katsikopoulos et al., 2020). Its detection sensitivity was 0.86 and the false-alarm rate 0.44. An “OR” classification rule (“If any of the indicators are present, classify as fake, otherwise non-fake”) had a higher sensitivity (0.97) but also a high false-alarm rate (0.66). The “email+hospital+citation” rule (Study 8) had greater sensitivity (0.90) and a smaller false-alarm rate (0.37).

Note that our tallying rule reflags likely fakes without certainty that a given publication is actually (legally) fake. Nevertheless, it is a reliable tool to red-flag scientific reports for further analysis and is a rational basis to estimate the upper value of fake publishing. Detecting fake papers with other indicators (e.g. image duplications, text plagiarism etc.) and in disciplines outside of biomedicine is expected to reveal different results.

### ESTIMATING THE INCIDENCE OF POTENTIAL FAKE PUBLICATIONS

When estimating the RFP incidence with our initial tallying rule (“email+NIA”) in the combined sample of Study 3 and 4, 589/4,001 RFP (15.7%) were found, where 328 (55.8%) were from China (Fig. 3; Tab. 4). Within-country percentages of RFPs vary considerably and are highest for Russia (48.3%), Turkey (47.5%), China (43.9%), Egypt (40.0%), and India (38.8%). However, when applying the validated “email+hospital” tallying rule, the RFP rate was 11% (110/1.000), with China contributing 79%, USA, Spain, Turkey and Korea 3-4%, and other countries 1% or less. This suggests that the main source of fake publications are Chinese hospitals.

**Figure 3.**
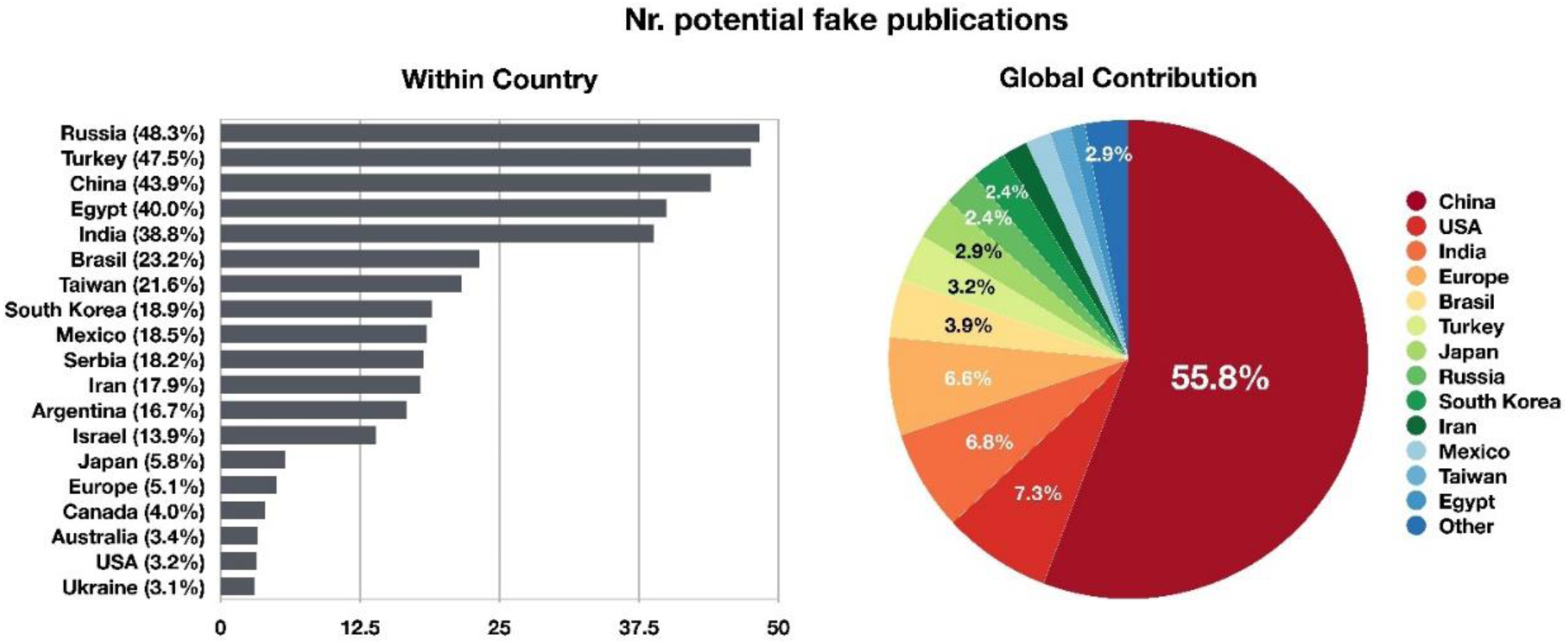
Estimating the global incidence of possible fakes in 2020. Left: Estimated percentage of red-flagged publications when applying the “email-NIA” indicator of each countrýs publication output (based on 2020 publications in Studies 3-5). There are three clusters: countries with high (>30%), medium (10%-30%), or low (<10%) numbers of RFPs. Interestingly, these clusters might reflect each natiońs average level of publication pressure: governmental (high), institutional (medium), or regular (low). Right: the contribution of different countries to the total number of potential fake publications (sample *N*=589). For absolute values, see Table 4. When applying the validated “email+hospital” tallying rule, the RFP contributions are: 79% China, 3-4% USA, Spain, Turkey, Korea; others 1% or less.

**Table 4.** Country analysis of potential fake publications (RFP)

| **Country** | **No. publications screened** | **No. RFP** | **% RFP within country** | **Global RFP number by country (%)** |
| --- | --- | --- | --- | --- |
| **Russia** | **29** | **14** | **48.3** | **2.4** |
| **Turkey** | **40** | **19** | **47.5** | **3.2** |
| **China** | **747** | **328** | **43.9** | **55.8** |
| **Egypt** | **15** | **6** | **40.0** | **1.0** |
| **India** | **103** | **40** | **38.8** | **6.8** |
| **Brazil** | **99** | **23** | **23.2** | **3.9** |
| **Taiwan** | **37** | **8** | **21.6** | **1.4** |
| **South Korea** | **74** | **14** | **18.9** | **2.4** |
| **Mexico** | **54** | **10** | **18.5** | **1.7** |
| **Serbia** | **11** | **2** | **18.2** | **0.3** |
| **Iran** | **56** | **10** | **17.9** | **1.7** |
| **Argentina** | **12** | **2** | **16.7** | **0.3** |
| **Israel** | **36** | **5** | **13.9** | **0.8** |
| **Japan** | **293** | **17** | **5.8** | **2.9** |
| **Europe** | **772** | **39** | **5.1** | **6.6** |
| Germany | 221 | 6 |  |  |
| Italy | 144 | 11 |  |  |
| Spain | 98 | 4 |  |  |
| France | 88 | 2 |  |  |
| Netherlands | 67 | 1 |  |  |
| Switzerland | 50 | 3 |  |  |
| Sweden | 45 | 1 |  |  |
| Poland | 32 | 4 |  |  |
| Greece | 15 | 4 |  |  |
| Portugal | 12 | 3 |  |  |
| **Canada** | **25** | **1** | **4.0** | **0.2** |
| **Australia** | **89** | **3** | **3.4** | **0.5** |
| **USA** | **1347** | **43** | **3.2** | **7.3** |
| **Ukraine** | **162** | **5** | **3.1** | **0.8** |
| **Sum** | **4001** | **589** |  | **100%** |

*Note:* Estimates of red-flagged (potential) fake publications (RFP), their rate by country (% RFP within country) and each country's contribution to the RFP pool (n=589) as a percentage. The table shows the percentage of publications (red-flagged/number screened publications) in Studies 1-6 (2020, neuroscience and medicine). Countries with <10 publications are not included in the table; they are Ireland (N=9/1), Pakistan (N=8/2), Thailand (N=6/2), Belarus (N=1/6), South Africa (N=5/2), Saudi Arabia (N=5/2), Czech Republic (N=3/1), Malaysia (N=3/1), Nigeria (N=3/1), Tunisia (N=3/2), Romania (N=2/1), and Morocco (N=1/1).

Given the 2020 global publication output of 1.33 million publications (Scimago; neuroscience/medicine) and an average of 11% RFPs in both fields using the validated tallying-rule, the annual RFP-incidence is nearly 150.000 in 2020.

Assuming that our RFP estimate reflects the true number of fake publications, and assuming an average $10,000 price tag for a fake publication order, the annual revenue of paper mills may be up to $1 billion. This revenue does not include non-Scimago journals (including “predatory journals”), nor the publication and open access fees charged by academic publishers (approx. $1 billion, similar to the estimated paper mill revenue). Although the incidence of true fake publications as identified by our detection criterion is expected to be smaller than our RFPs if the number of false alarms exceeds that of missed actual fakes, the estimate of an annual 150.000 potential fake publication rate is worrying, and it is on the rise.

### QUALITATIVE ANALYSIS OF PAPER MILL STRATEGIES

More than 1,000 paper mills openly advertise their services on Baidu and Google to “help prepare” academic term papers, dissertations, and articles intended for SCI publications. Most paper mills are located in China, India, UK, and USA, and some are multinational. They use sophisticated, state-of-the-art AI-supported text generation, data and statistical manipulation and fabrication technologies, image and text pirating, and gift or purchased authorships. Paper mills fully prepare – and some guarantee –publication in an SCI journal and charge hefty fees ($1,000-$25,000; in Russia: $5,000) (Chawla, 2022) depending on the specific services ordered (topic, impact factor of target journal, with/without faking data by fake “experimentation”). An unsolicited meeting with a paper mill provided a rare and authentic inside view of their business practices (Tab.1).

Paper mills employ science graduates, academicians, and (sometimes naïve) scientific consultants for editorial help who work in countries with high English aptitude (UK, USA, India). They also offer “rewards” (bribes) to editors for publishing their fabrications (Tab.1). We know of at least 12 such cases (two reported by editors, 10 acknowledged by an academic publisher who asked not to be identified). Here, editors were offered payment for each publication and were lured by a “citation booster” whereby paper mills offered to cite the “friendly” journal in their other fake articles. Although we do not know how many editors have received or accepted such bribes, it is an unprecedented and disturbing fraud-for-profit corruption case of scholarly publishing.

## DISCUSSION

The dramatic rise of fake science publishing is driven by an unscrupulously corrupt – and increasingly successful – paper mill industry responsible for an estimated 150,000 RFPs annually as of 2020, a number considerably higher than current estimates (Tab.2). Our 2020 estimate of 11% RFPs much higher than the 2011 estimate of 0.1% fake publications in China (Hu and Wu, 2013) and the 1% listed in Tab.1, close to the 5%-10% reported in a pharmacology (Seifert, 2021) and a cancer journal (Heck et al., 2021), but below the 21-32% “honorary” and ghost authorship cases reported in biomedicine (Flanagin et al., 1998; Wislar et al., 2011). By now, paper mills have likely emerged as a billion-dollar global industry, magnitudes higher than the $4.5 million monetary value estimated in 2011 (Hu and Wu, 2013).

Fake science publishing is known to originate mainly from China (Hu and Wu, 2013; Lei and Zhang, 2018; Mallapaty, 2020; Schneider, 2021), India (Elango, 2021), and Russia (Abalkina, 2021), and, as we showed, it has now evolved into a rapidly growing industry of fake science publishing. Our analysis confirms the existence, continuous growth, and notable scope of fake publishing, with most red-flagged publications coming from Chinese hospitals (79% in 2020).

The rapid rise of the fake science industry is driven by SCI publication pressure on medical doctors and scientists, who hire paper millś ghostwriting services at $1,000 to 25,000 per publication (Hu & Wu, 2013). Academic publishers have acknowledged the problem and recently implemented a detection tool (Else, 2022; see also COPE & STM Committee on Publication Ethics, 2022). It remains to be seen how effective this tool will be, how diligent and transparent the academic publisherś effort is to filter out submitted fake manuscripts and remove published ones from the permanent scientific record. Chinese authorities, although aware of the situation (Cyranoski, 2018), have not resolved the problem. If quantity of publication output is viewed as an index for becoming the world leader in science, then paper mills contribute to reaching this goal. Applying this metric, China has almost caught up with the US in publication output in only one decade (www.nature.com/nature-index/country-territory-research-output). We submit that it should be in their best interest to remove the publication pressure on their hospitals to prevent the knowledge pollution of the scientific record.

Paper mills feed on the rising administrative practice throughout the world to evaluate researchers mainly by the “publish-or-perish“ criterion of counting papers and journal impact factors as a surrogate for evaluating actual research quality and content (Van Dalen and Henkens, 2012; Candal-Pedreira et al., 2022).

Fake academic publishing is a major driver of global science publishing growth and is a considerable and growing risk for medical practice. For example, Byrne showed that 712 problematic papers were cited >17,000 times and estimated that about one quarter of them may misinform future development of human therapies (Park et al., 2022). Preclinical studies at biotech company Amgen could replicate the results of only 6/53 “landmark” articles, and at Bayer, only 14/67 were replicable in oncology, women’s health, and cardiovascular medicine. The “replication crisis” slows down the development of life-saving therapies with an estimated financial loss of $28 billion annually by the pharmaceutical industry (Gigerenzer, 2018). Yet another example of how scientific fraud can affect medical practice is reported by Avenell et al. (2019). After assessing the citations of 12 retracted clinical trial reports in 68 systematic reviews, meta-analyses, guidelines, and clinical trials, they concluded that 13 out of the 68 reviews would likely have to change their conclusions if the retracted publications were removed.

It is important to keep in mind that our indicators provide a red flag, not legal proof, that a given manuscript or publication might be fake. However, it is the authorś – not the editors’ burden of proof to demonstrate that their science can be trusted. Whether this type of scientific misconduct is a conspiracy to commit injurious falsehood or a crime is for others to decide.

Industrial-style fake science publishing is possibly the biggest science scam of all times, wasting financial resources, slowing down medical progress, and possibly endangering lives. The damage already done is unknown, and a realistic impact assessment of fake science is not yet available. The emergence of Chat-GPT and other sophisticated large language models will amplify the problem, as they lower the production cost of fake papers. Our easy to apply indicators can be a first step to help stave off fake publishing.

Halting this development requires an immediate response. But what can be done? First, our simple detection tallying method can be used by reviewers and editors to red-flag potential fakes with or without additional indicators. Second, the academic community should consider revising its practice to judge scientistś productivity mostly (or solely) on surrogate quantitative criteria (publication numbers, h-factors, impact factors, etc.) and instead evaluate the content, quality and relevance of their research (Van Dalen and Henkens, 2012). The European Research Council (ERC) has already taken a first step by asking researchers to refrain from listing impact factors in their applications, consistent with the San Francisco Declaration on Research Assessment (DORA). Thirdly, we need an advanced system to check scientific integrity which is independent of academic publishers. Finally, learned societies, funding agencies, and governmental bodies should consider sanctioning fake polluted journals and their publishers.

Until science publishing fraud is largely eradicated, the collateral damage of fake science poses the risk that scientific analyses, experiments, and clinical trials will more likely fail, public health information will be less accurate or (intentionally) misleading, and presumably effective and safe therapies may not deliver what was promised. It also runs the risk that the public loses its trust in the honesty of science itself. Simple detection of fake publications, as proposed here, or more complex automated methods can help prevent further damage to the permanent scientific record and enable the retraction of fake publications at scale. We propose a “call to action” to restore the integrity of our global knowledge base in biomedicine, science, and technology.

## Data Availability

All data produced in the present study are available upon reasonable request to the authors

## Notes

### Competing Interest Statement

The authors have declared no competing interest.

### Funding Statement

The study was funded by the Otto-v.-Guericke University of Magdeburg and by a grant from the State of Sachsen-Anhalt Ministery of Science, Magdeburg, Germany

### Summary of Updates

In this revision we clarified which tallying rule was used for each experiment. In addition, we calculated the red flag incidence with the validated "institutional email+hospital" indicator and adjusted the total estimate of red flagged publications. We appreciate the suggestion of the readers suggesting it and clarified this in the text after analyzing the yr 2020 sample of the data again to obtain more valid estimate of the scope of fake publishing. The result does not change the overall conclusion of our paper

